# Growth Prediction of Type B Aortic Dissections Using Wall-Stress-Driven Finite Element Simulation Based on the Unified-Fiber-Distribution (UFD) Model

**DOI:** 10.64898/2025.11.24.25339532

**Authors:** Xue Liang, Marc-Philipp H. Schmid, Minliang Liu, Hannah L. Cebull, Michael Zhang, Sunny Xu, Muhammad Naeem, John N. Oshinski, John A. Elefteriades, Rudolph L. Gleason, Bradley G. Leshnower, Hai Dong

## Abstract

Type B aortic dissection (TBAD) is a serious, potentially life-threatening condition which occurs when a tear develops in the inner lining (intimal layer) of the descending aorta, causing the layers of the aortic wall to separate (dissect) and creating true and false lumens. TBAD can be classified into complicated and uncomplicated types based on the presence of complications (e.g., rupture or malperfusion). For complicated TBAD, the standard treatment is thoracic endovascular aortic repair (TEVAR) with a stent graft. Uncomplicated TBAD can be managed with optimal medical therapy (OMT). Predicting growth and aneurysmal progression of uncomplicated TBAD is clinically important for timing of intervention during OMT. In this study, we extended our previously developed finite element (FE)-based tissue growth framework and applied it to predict the precise geometry and diameter growth of TBAD. Specifically, the unified-fiber-distribution (UFD) model was applied to describe aortic wall mechanics, and a novel centerline-based algorithm was developed to determine the local material coordinates of aortic tissues. A linear kinematic growth law related to local wall stress was used for tissue growth. Patient-specific aortic geometries from three serial computed tomography (CT) scans were obtained for seven patients with TBAD. Using the first two CT images and each patient’s blood pressure, inverse FE analysis was performed to obtain patient-specific growth parameters. These parameters were then used to simulate forward growth and predict geometry at the third time point. Predicted aortic geometries and dimensions matched well with *in vivo* measurements: across all patients the maximum diameter error was below 3.5% and the mean diameter error below 4%. Such accurate patient-specific growth forecasts demonstrate the potential of this computational framework to support clinical decision-making in uncomplicated TBAD.

## 1. Introduction

Type B aortic dissection (TBAD) is a life-threatening event in which a tear in the descending aortic intima allows pressurized blood to split the aortic wall into a true lumen and a pressurized false lumen [1]. By Stanford classification, TBAD involves the descending thoracic aorta distal to the left subclavian, unlike type A dissections of the ascending aorta. Clinically, TBAD is subdivided into complicated and uncomplicated cases based on the presence of complications (e.g., rupture or malperfusion). Complicated TBAD generally requires urgent repair – typically thoracic endovascular aortic repair (TEVAR) with a stent graft [2] – whereas uncomplicated TBAD is managed initially with optimal medical therapy (OMT) which primarily involves anti-hypertensive therapy and surveillance imaging [3]. However, even under OMT many patients with TBAD could experience progressive aneurysmal degeneration of the false lumen [4–6], which underscores the clinical importance of tracking TBAD geometry: early identification of rapid enlargement could prompt prophylactic intervention before rupture.

Following dissection, the aortic diameter could increase substantially. For example, Rylski et al. [7] reported that the proximal descending aorta’s diameter grows as many as ∼6.4 mm (≈23%) immediately after a type B dissection. During chronic follow-up, a large fraction of TBAD patients exhibit further expansion: Sueyoshi et al. [8] found that roughly 75% of aortic segments in chronic TBAD enlarge over time, with mean growth rates on the order of several millimeters per year. Such enlargement carries risk: even moderate increases in aortic diameter amplify the chances of rupture or malperfusion. Thus, accurately predicting TBAD progression – from serial imaging or other data – has clear value. A validated growth forecast could guide the timing of interventions (such as elective TEVAR) during OMT, when the aorta reaches a high-risk size or growth rate.

Biomechanical modeling has the potential to inform such predictions, but existing computational studies on TBAD have largely focused on flow dynamics rather than tissue growth. For instance, computational fluid dynamics (CFD) and 4D-flow MRI analyses have revealed that complex hemodynamic patterns (high flow helicity, low oscillatory wall shear) correlate with false-lumen aneurysmal expansion [1]. Fluid–structure interaction (FSI) simulations have explored how flap motion and wall compliance affect thrombus formation in the false lumen [9–11]. These studies highlight the role of hemodynamics in disease progression, but they do not directly forecast changes in vessel geometry. Growth and remodeling of the aortic wall involve biological adaptation to loading over weeks or months, which calls for constitutive and growth models.

In our prior work [12], we introduced a finite-element (FE)-based framework for aortic-root aneurysm growth after V-shape repair [13, 14]. In this work, we extended this finite element (FE)-based tissue growth framework and applied it to predict the precise geometry and diameter growth of TBAD. Specifically, the unified-fiber-distribution (UFD) model [15–19] was applied to describe aortic wall mechanics, and a novel centerline-based algorithm was developed to determine the material coordinates of aortic tissues. The kinematic growth law related to local wall stress was used for tissue growth. Patient-specific aortic geometries from three serial computed tomography (CT) scans were obtained for seven patients with TBAD. Using the first two CT images and each patient’s blood pressure, inverse FE analysis was performed to obtain patient-specific growth parameters. These parameters were then used to simulate forward growth and predict geometry at the third time point. Predicted aortic geometries and dimensions matched well with *in vivo* measurements: across all patients the maximum diameter error was below 3.5% and the mean error below 4%. Such accurate patient-specific growth forecasts demonstrate the potential of this computational framework to support clinical decision-making in uncomplicated TBAD.

## 2. Methods

### 2.1 Patient data collection

We retrospectively collected clinical CT images from seven TBAD patients (P1–P7 summarized in Table 1) who underwent open aortic replacement at Emory University Hospital (Atlanta, Georgia). For each patient, CT images at three preoperative follow-up time points were available. The in-plane spatial resolution of the CT images ranged from 0.69 × 0.69 mm to 1.07 × 1.07 mm, with slice thicknesses of 1.00–2.00 mm. Patient-specific imaging parameters are provided in Appendix A. All participants gave written informed consent, and the study was approved by the Emory University Institutional Review Board (IRB00109646 and STUDY00002737).

**Table 1.** Patient demographics: Gender, systolic pressure (𝑃_sys_), diastolic pressure (𝑃_dia_), and the time intervals ΔT_12_(CT-1 to CT-2) and ΔT_23_(CT-2 to CT-3).

| Patient ID | P1 | P2 | P3 | P4 | P5 | P6 | P7 |
| --- | --- | --- | --- | --- | --- | --- | --- |
| Gender | M | M | M | M | M | F | F |
| $P_{sys}$ (mmHg) | 161 | 155 | 120 | 180 | 137 | 162 | 191 |
| $P_{dia}$ (mmHg) | 91 | 94 | 72 | 99 | 70 | 84 | 94 |
| $\Delta T_{12}$ (month) | 75.5 | 20 | 18.5 | 25 | 42 | 75.5 | 30.5 |
| $\Delta T_{23}$ (month) | 39 | 12.5 | 29 | 48 | 20.5 | 25.5 | 24.5 |

### 2.2 Construction of Patient-Specific Finite Element Model

Based on the CT images (e.g., Fig. 1a), patient-specific geometries of the TBAD (Fig. 1b) were segmented using 3D Slicer (www.slicer.org) for each patient at each of the three time points. The three-dimensional (3D) aortic surface of the TBAD outer wall was then reconstructed in 3D Slicer, exported, and imported into Altair HyperMesh 2021 (Altair Engineering, Inc.) to generate the finite element mesh for the descending aorta of the outer wall (Fig. 1c). We first created 4-node quadrilateral shell elements (S4) with an element size of approximately 1.5 × 1.5 mm, based on the reconstructed surface geometry. Four layers of eight-node linear brick elements (C3D8) were generated by offsetting the S4 elements [20, 21]. A uniform thickness of 2 mm was assumed for the aortic wall thickness. Finite-element (FE) analyses and the optimization procedure were performed using Abaqus 2024 and MATLAB R2024a, respectively.

**Fig 1.**
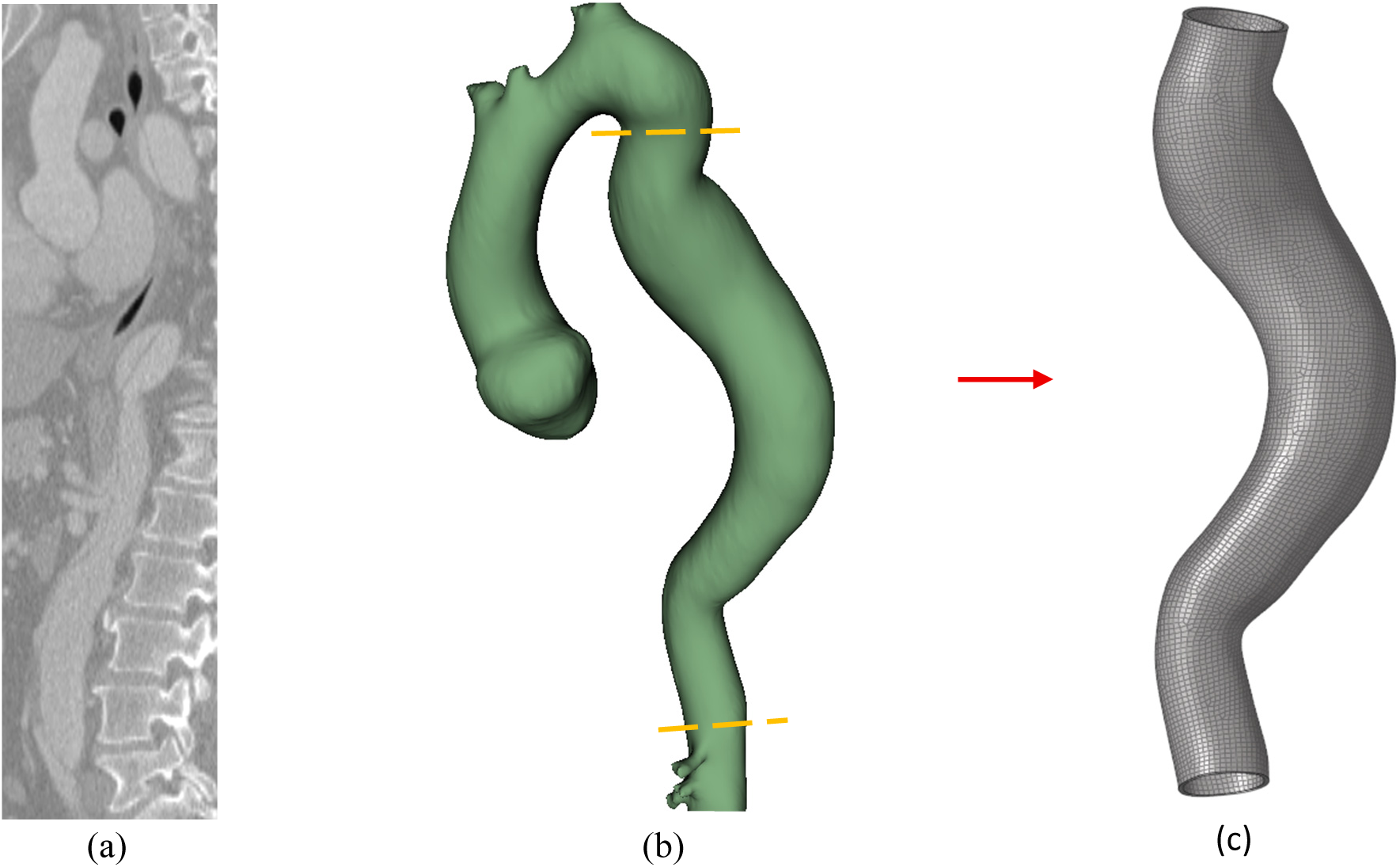
(a) Preoperative CT image of the aorta from a representative patient (P1); (b) 3D aortic surface reconstructed from (a) in 3D Slicer; (c) finite-element mesh generated in HyperMesh 2021 (Altair Engineering) from the geometry in (b), with trimmed boundaries.

### 2.3 The Growth Framework based on the Unified-Fiber-Distribution Model

The total deformation gradient of the tissue can be decomposed multiplicatively into elastic and growth parts [12, 22],

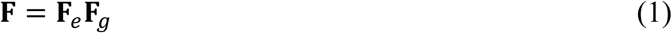

where 𝐅_e_is the elastic deformation gradient mapping the current reference state to the current loaded state under mechanical loading. The constitutive relation between the applied loading and 𝐅_e_ was characterized by our recently developed unified-fiber-distribution (UFD) model [19]. 𝐅_g_ is the growth deformation gradient mapping the original reference state to the current (grown) reference state. The kinematic growth law (Sec. 2.3.2) related to local wall stress was used for tissue growth.

#### 2.3.1 Unified-Fiber-Distribution (UFD) Hyperealstic Model

Previously, we developed a unified-fiber-distribution (UFD) model [19] that treats collagen fibers as a unified distribution rather than as two or several discrete fiber families, as in the widely used Gasser–Ogden–Holzapfel (GOH) model [23]. In this framework, neither the specific distribution form nor the number of fiber family needs to be prescribed. By representing fibers as a unified distribution, the UFD model is expected to be more consistent with the actual fiber architecture of aortic tissue. Here, we used the UFD model to characterize the hyperelastic behavior of aortic tissue.

The total strain-energy function can be additively decomposed into isochoric and volumetric parts.

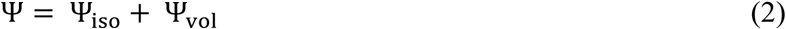

where the volumetric part is Ψ_vol_ = ^1^ (𝐽− 1)^2^, with 𝐷 a material (penalty) parameter enforcing near-incompressibility under the assumption that aortic tissue is approximately incompressible. The isochoric part can then be written as:

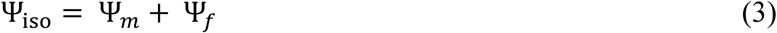

where Ψ_m_ denotes the matrix contribution to the strain energy, described by the neo-Hookean model [24–28].

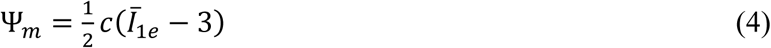

where 𝑐 is the initial shear modulus of the matrix, 𝐼_1̅e_ = tr(𝐂^-^_e_) is the first invariant of 𝐂^-^_e_ = 𝐽^-2/3^𝐂_e_, with 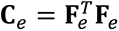 denoting the right Cauchy–Green tensor and 𝐽_e_ = det𝐅_e_ > 0 is the determinant of the elastic deformation gradient 𝐅_e_. The term Ψ_f_denotes the fiber contribution to the strain energy—which distinguishes the UFD model from other formulations—and is given in [19].

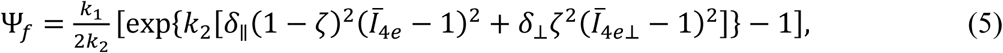

where 𝐼̅_4e_ = (𝐚_O_⊗𝐚_O_): 𝐂^-^_e_ and 𝐼̅_4e..l_ = (𝐚_O..l_⊗𝐚_O..l_): 𝐂^-^_e_, with 𝐚_O_ and 𝐚_O..l_ denoting unit vectors in the circumferential (mean-fiber) and axial (cross-mean-fiber) directions, respectively. The parameters 𝑘_1_, 𝑘_2_ and 𝜁 are fiber-related material constants: 𝑘_1_ represents the initial stiffness, 𝑘_2_ governs strain-stiffening under stretch, and 𝜁 is a scalar that characterizes the fiber distribution in an integral sense. The indicators 𝛿_∥_ and 𝛿_⊥_ exclude contributions from fibers in compression, implemented via the Heaviside step function.

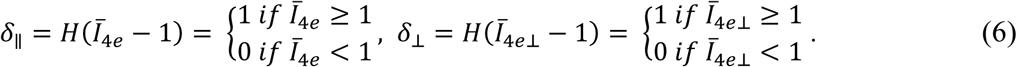

The material parameters for the UFD model in this study are listed in Table 2.

**Table 2.** The parameters used in the UFD model [19] are listed; 𝑅^2^ describes the goodness of fit to the experimental data [29].

| Parameters | $c$ (kPa) | $k_1$(kPa) | $k_2$ (-) | $\zeta$(-) | $D$ (kPa<sup>-1</sup>) | $R^2$ |
| --- | --- | --- | --- | --- | --- | --- |
| Values | 222.07 | 101.81 | 99.99 | 0.46 | $1 \times 10^{-6}$ | 0.93 |

#### 2.3.2 Growth Evolution Law

We assume that the growth deformation 𝐅_g_ in Eq. (1) can be expressed as:

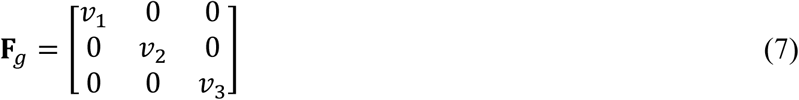

where 𝑣_1_, 𝑣_2_ and 𝑣_3_ are the growth stretch multipliers. The directions {1, 2, 3} the circumferential, axial, and radial directions, respectively.

Aortic wall stress has been shown to be strongly associated with aneurysm dilation [30, 31]. With this in mind, we assume the evolution of the growth rate 𝑣·_1_ depends linearly on 𝜎_1_ (the maximum principal Cauchy stress), and the evolution of 𝑣·_2_depends linearly on 𝜎_2_(the intermediate principal Cauchy stress). As the wall thickness is unavailable, we assume it remains unchanged between CT-1 and CT-2. The growth evolution laws are thus:

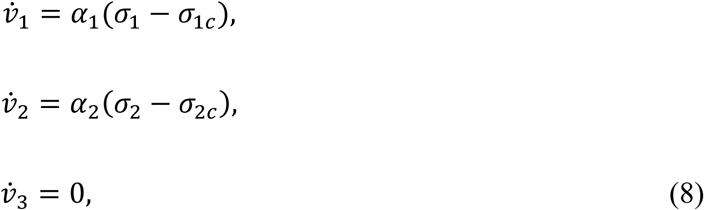

where {𝛼_i_, 𝜎_ic_} (𝑖 = 1, 2) are growth parameters. The thresholds 𝜎_ic_ (𝑖 = 1, 2) > 0 can be interpreted as critical trigger stresses for growth: growth occurs when 𝜎_i_ > 𝜎_ic_ and is absent when 𝜎_i_ < 𝜎_ic_. The effective growth-driving stress is then the true stress minus the critical trigger stress. During the simulation, the principal stress components were obtained via spectral decomposition of the Cauchy stress tensor; the three eigenvalues correspond to the three principal stresses.

The UFD model and the growth evolution law were implemented [12] in the UMAT user-defined material subroutine of Abaqus/Standard (SIMULIA, Providence, RI). A numerical approximation scheme [32, 33] was used to calculate the tangent modulus tensor.

### 2.3 A Novel Centerline-Based Algorithm to Assign the Local Material Coordinate Systems

A centerline-based algorithm was developed and implemented using MATLAB R2024a to assign local coordinate systems. First, the inner surface was read from the input file and the aortic centerline—semi-automatically created on the 3D aortic reconstruction to represent the medial axis—was imported. This produced four arrays: two storing node IDs and coordinates, and two storing element IDs and their node connectivity (or centerline connectivity). For each inner-surface element, the following steps were performed. As in Abaqus, the 3-direction (𝒆_3_) was defined as the inward unit normal to the element. The element centroid was then computed from its nodes, and the nearest centerline node to this centroid was identified; together with its preceding centerline node, it defined a centerline segment. A plane 𝑃_intersect_containing this centerline segment and 𝒆_3_ was constructed. The intersection of 𝑃_intersect_ with the element plane defined the 1-direction; normalizing this intersection vector yielded 𝒆_1_ . The projection of the centerline segment onto the element surface via the two intersecting planes is illustrated in Fig. 2. The remaining basis vector 𝒆_2_ was obtained from 𝒆_1_ × 𝒆_3_. The directions 𝒆_1_, 𝒆_2_ and 𝒆_3_ correspond approximately to the axial, circumferential, and radial directions, respectively. Because the four aortic-wall layers in the FE model were generated by offsetting, the local material coordinate systems determined on the inner surface were transferred to the elements of each layer. The pseudocode for this algorithm is summarized in **Table 1**.

**Fig 2.**
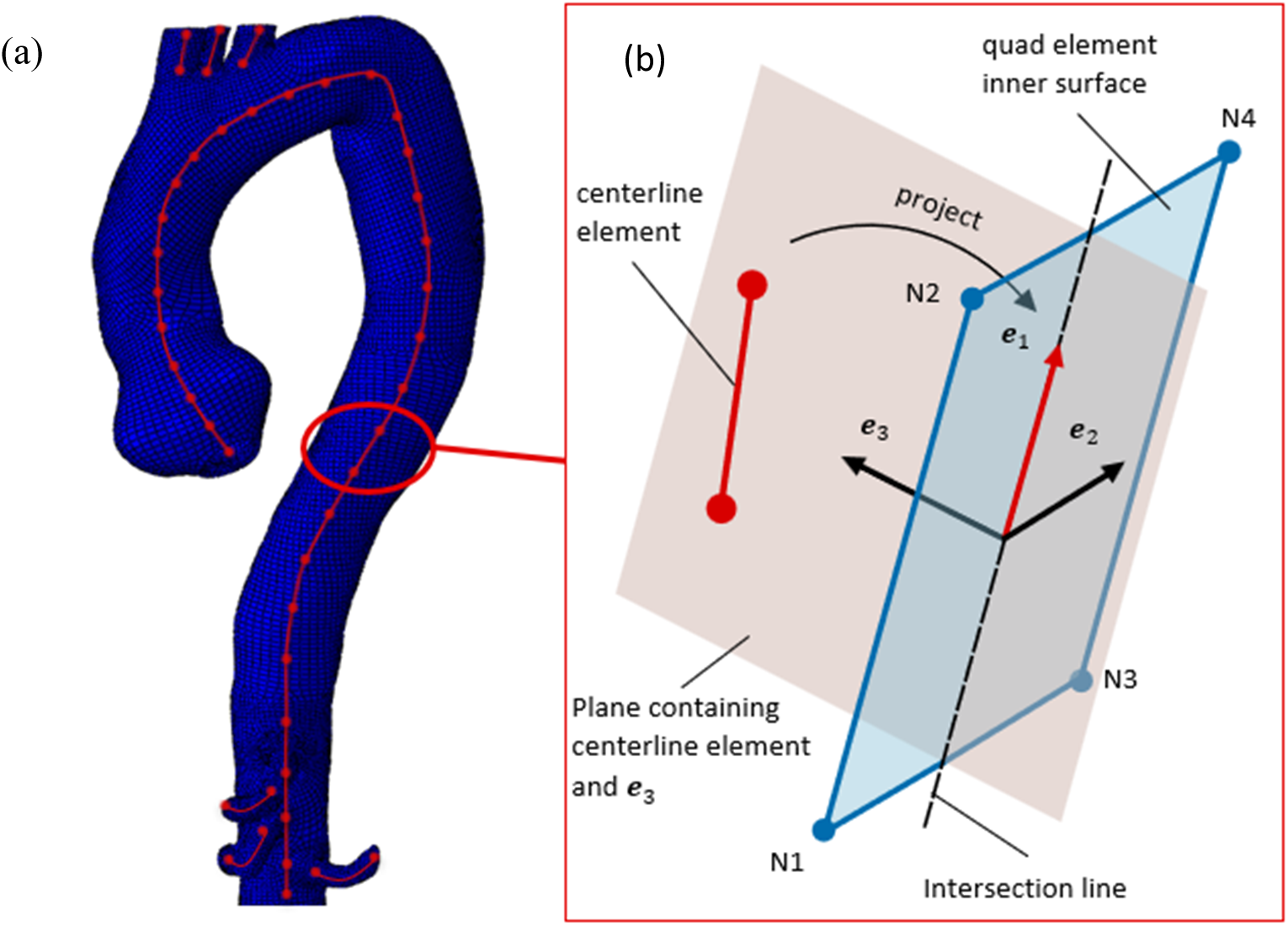
Illustration of the centerline-based algorithm to assign local coordinate systems for each finite element. 𝒆_1_, 𝒆_2_ and 𝒆_3_ are basis vectors of local coordinate system. 𝑁_1_, 𝑁_2_, 𝑁_3_, 𝑁_4_ are four nodes of quad element in inner surface. (a) Excerpt of the aortic model with a schematic centerline; (b) two planes whose intersection defines the 1-direction.

**Table 1.**
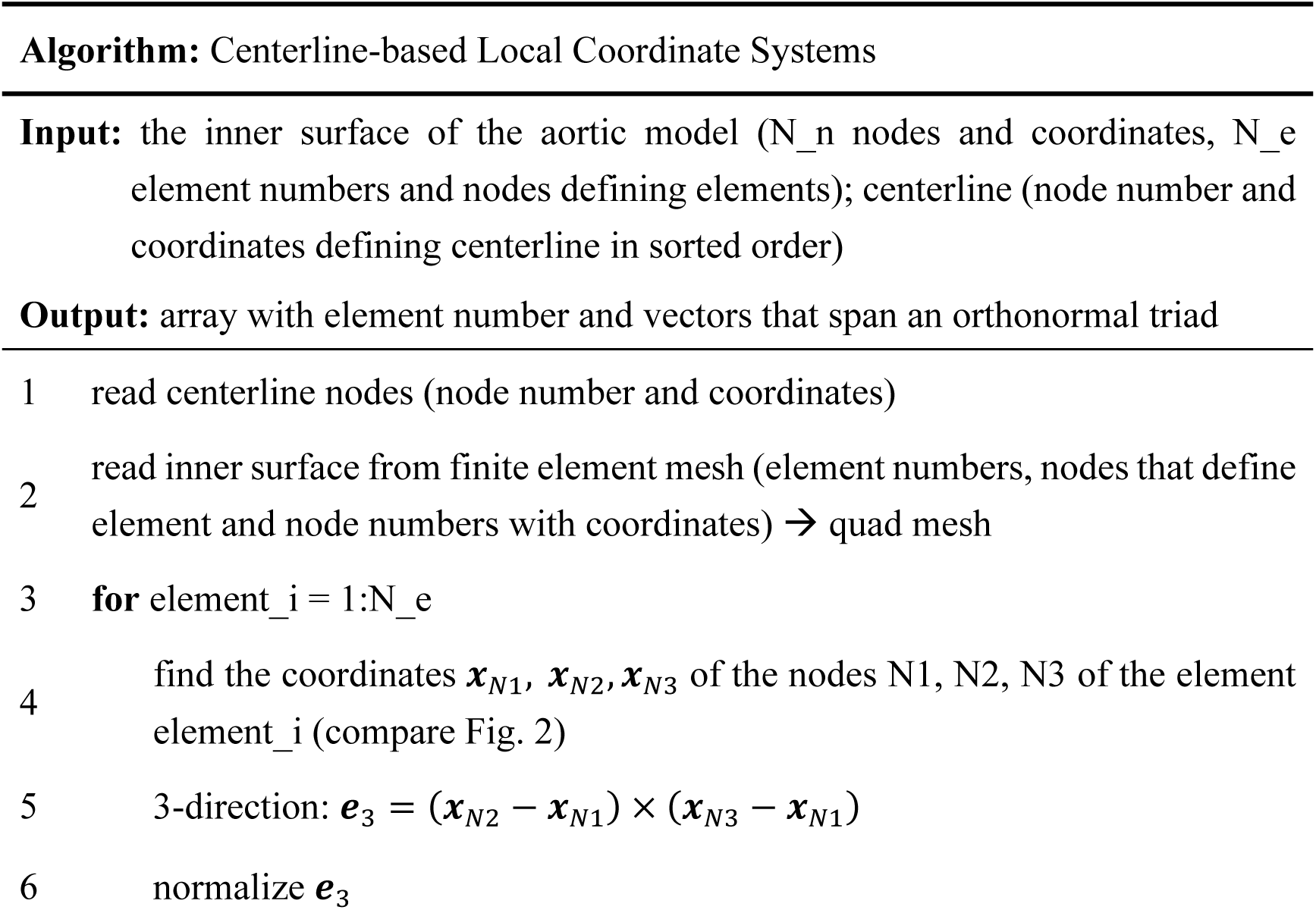
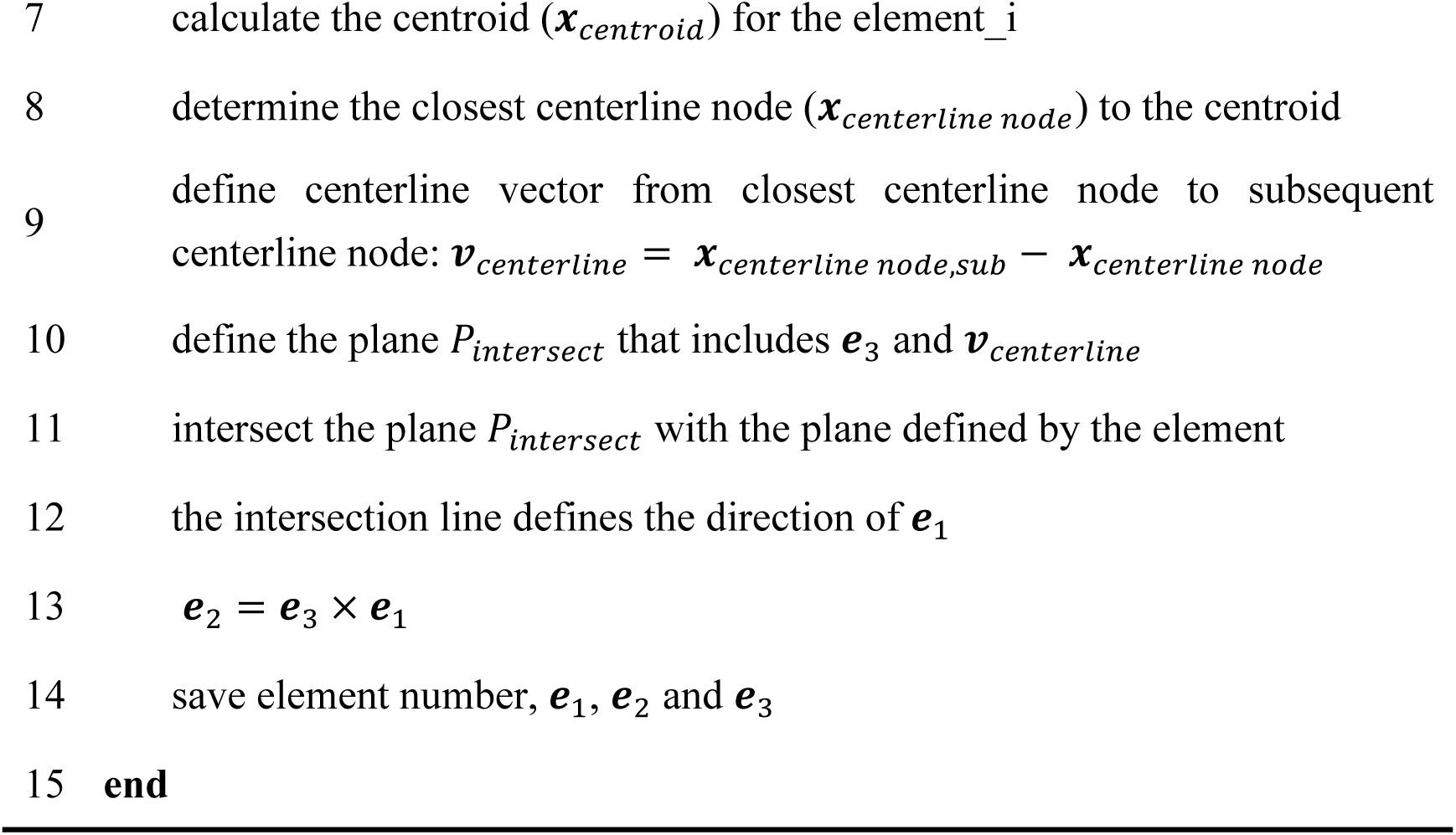
Centerline-based algorithm as pseudocode.

### 2.4 Growth Simulation and Optimization of Growth Parameters

We simulated patient-specific growth of the descending aorta starting from the CT-1 geometry (Section A, Fig. 3). Because this geometry reflects the *in-vivo* configuration under systolic pressure, directly applying physiological systolic pressure to it would yield inaccurate strains and stresses. Instead, the FE simulation should begin from the zero-pressure configuration. We therefore used the improved backward displacement method [34] to recover the zero-pressure geometry of the CT-1 aorta (Section A, Fig. 3). The patient-specific systolic pressure was then applied to the inner surface of this zero-pressure configuration to obtain the stress and strain fields in the deformed state (Section B, Fig. 3).

**Fig 3.**
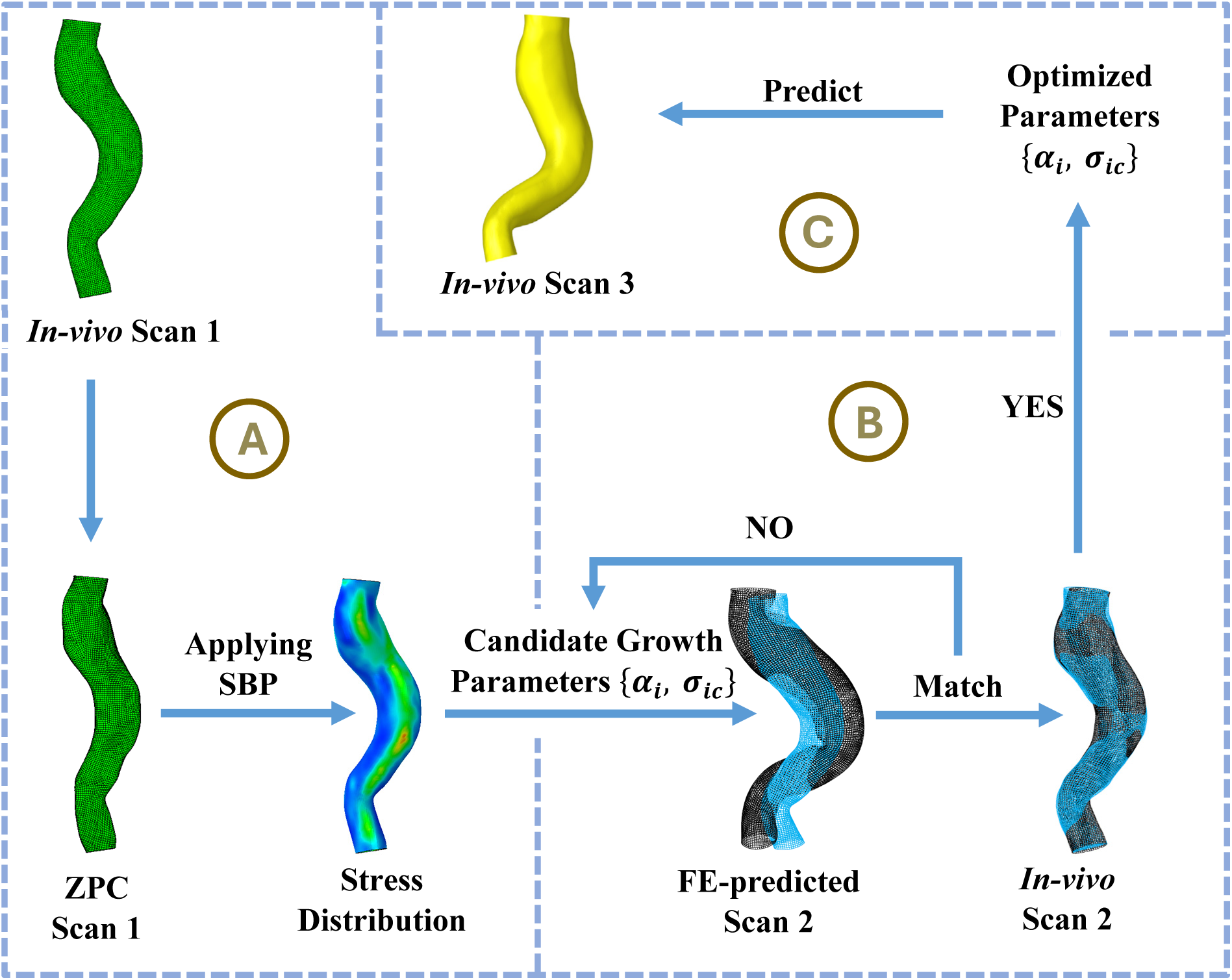
Overview of the optimization procedure.

Next, growth was initiated with a set of candidate parameters, and the growth stretches were computed from the laws in Eq. (8) using the stress field from Section A in Fig. 3. The process was terminated when the elapsed growth time reached CT-2, yielding the FE-predicted CT-2 configuration of the descending aorta. The parameters {𝛼_i_, 𝜎_ic_} were then tuned until the FE-predicted inner surface at CT-2 matched the *in-vivo* inner surface (Section B in Fig. 3). During the simulation, circumferential and axial displacements were fixed at the boundary of the upper and lower sections, while radial displacement at the boundaries was left unconstrained [35].

Specifically, we applied a least-squares procedure to match the FE-predicted and *in-vivo* inner surfaces of the descending aorta at CT-2. We define the node-to-surface distance between a mesh node 𝐗 on the predicted inner surface and the triangular mesh Ω of the *in-vivo* inner surface at CT-2 as [36]:

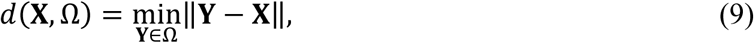

where 𝐗 is the coordinate vector of the node, 𝐘 is the coordinate vector of a node on the surface mesh Ω, and ‖𝐙‖ denotes the 3D Euclidean norm of an arbitrary vector 𝐙. The value 𝑑(𝐗, Ω) converges to a fixed limit as the mesh of Ω is refined (i.e., as element size decreases). In this study, the distance-convergent mesh size for Ω was approximately 0.5 mm. The growth parameters

{𝛼_i_, 𝜎_ic_} were then optimized by minimizing the sum of squared node-to-surface distances between the FE-predicted and *in-vivo* inner surfaces of the descending aorta.

We implemented the optimization using the *lsqnonlin* function in MATLAB 2024a (MathWorks, Natick, MA). Finite element simulations were run in Abaqus/Standard 2024 (SIMULIA, Providence, RI) on a Dell precision 3680 workstation (Intel Core i7-14700K @ 3.4 GHz, 20 cores/28 threads, 64 GB RAM, 64-bit).

Fig. 4 compares the CT configurations (CT-1, CT-2, and CT-3) at three pre-surgical follow-up time points for a representative patient (P1). To capture the relative position of a node on the CT-1 inner surface with respect to the CT-2/CT-3 inner surfaces, we define a signed distance parameter as:

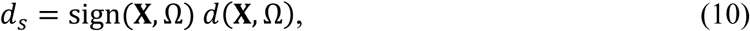

where 𝑑(𝐗, Ω) is given in Eq. (9), with sign(𝐗, Ω) = 1 when 𝐗 lies outside the surface Ω and sign(𝐗, Ω) = −1 when 𝐗 lies inside Ω.

**Fig 4.**
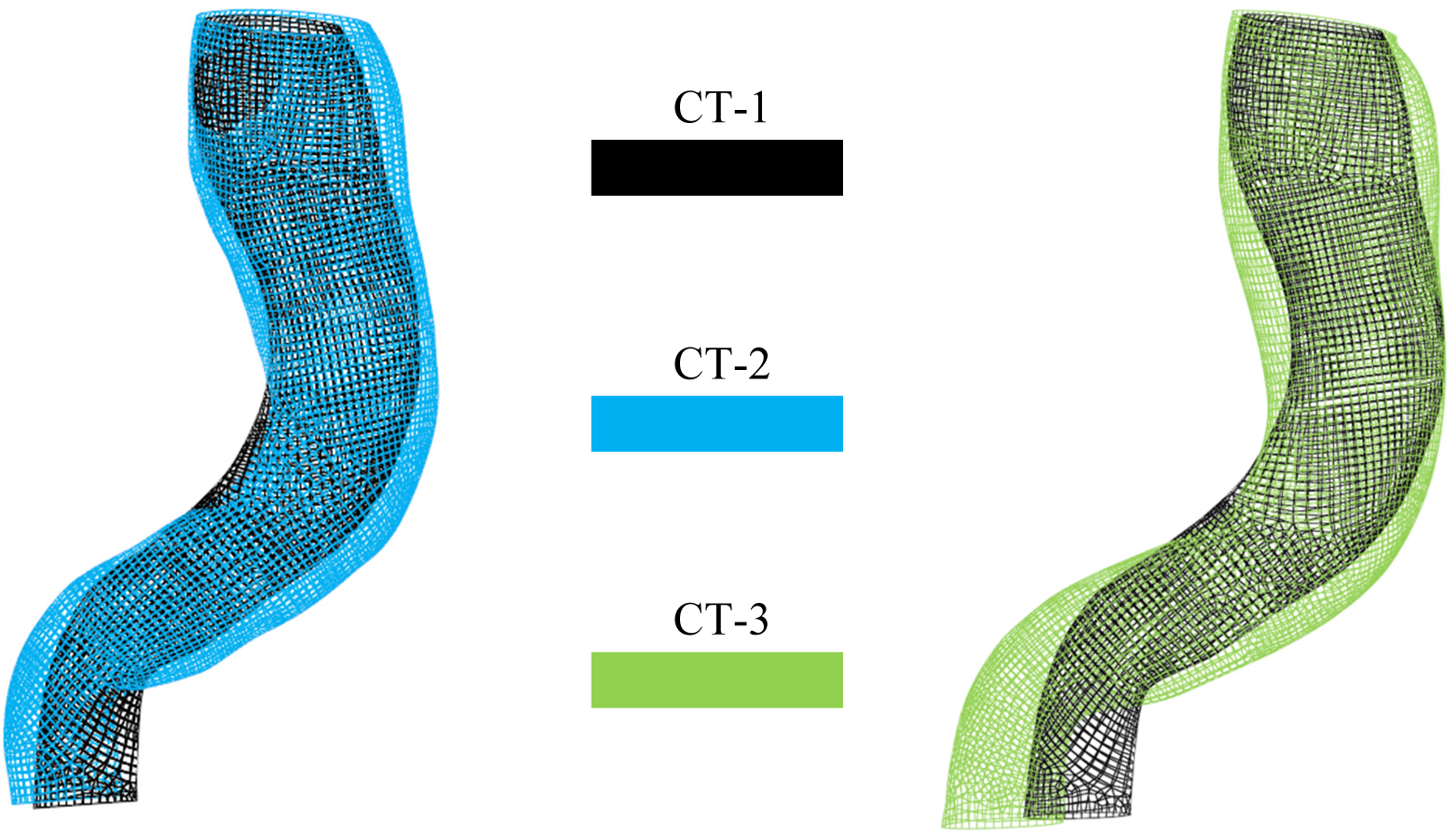
*In-vivo* geometries of the descending aorta for a representative patient (P1) at three preoperative follow-up time points (CT-1, CT-2 and CT-3), reconstructed from clinical CT images.

### 2.5 Diameter calculation

For the predicted configuration (CT-2-Growth) and the *in-vivo* configuration (CT-3) of each patient, diameters at 50 locations along the descending thoracic aorta, evenly spaced along the centerline (Fig. 5a), were extracted using custom MATLAB scripts. Based on these 50 measurements for CT-2-Growth and CT-3, the mean and maximum diameters of the descending thoracic aorta were computed

**Fig 5.**
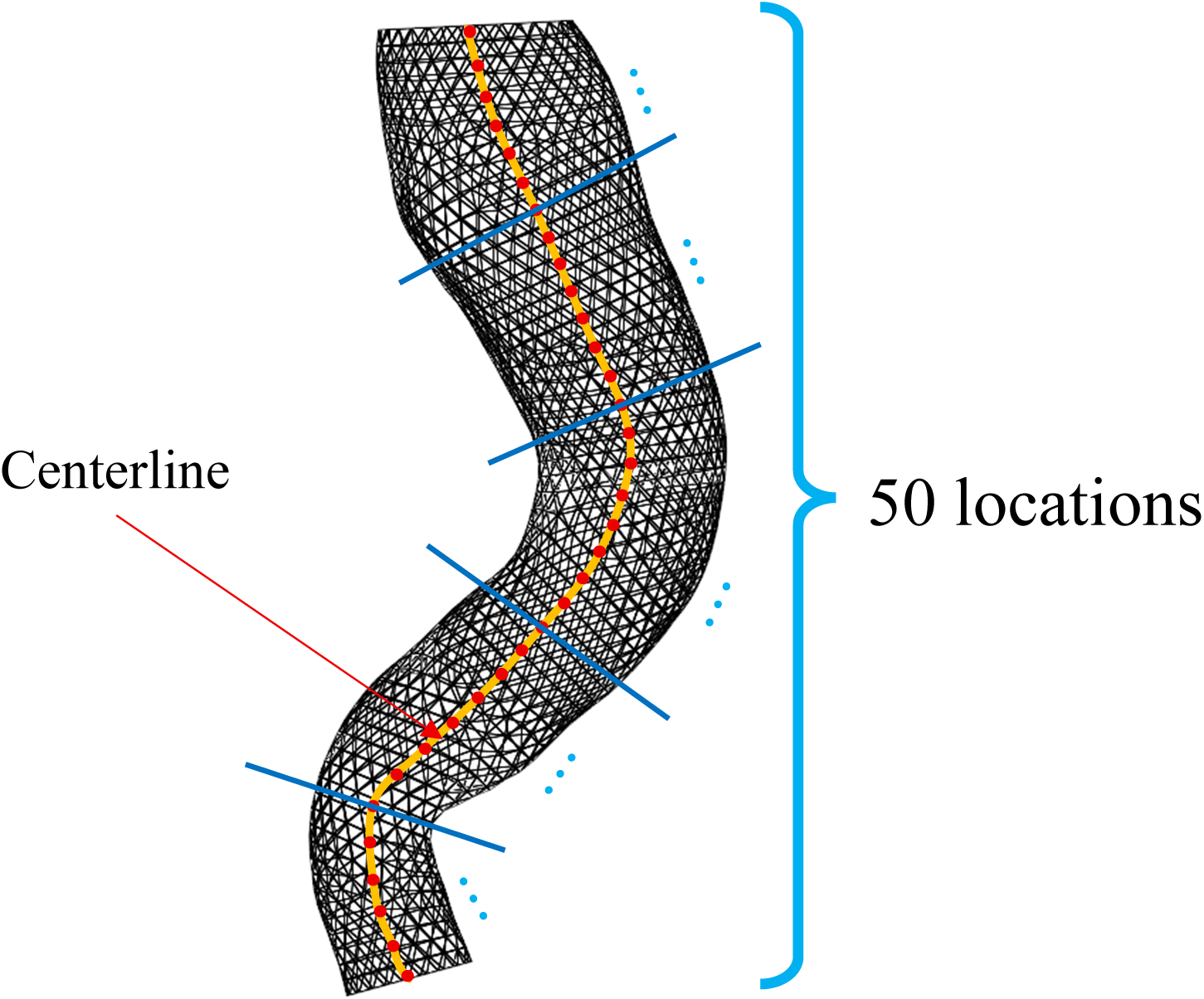
Triangular mesh and 50 sampling locations for a representative patient (P1).

## 3. Results

### 3.1 Optimization Process and Estimated Growth Parameters

Figure 6a compares the FE-predicted CT-2 geometry (growth from CT-1, denoted CT-1-Growth) at the first iteration with the CT-2 *in-vivo* geometry for P1. The corresponding *in-vivo* CT-1 and CT-2 geometries are shown in Fig. 4. The initial result indicates overpredicted growth: the mean node-to-surface distance between CT-1-Growth and CT-2 was +14.16 mm, implying that the predicted surface lay predominantly outside the *in-vivo* CT-2 surface. After iterative tuning of the candidate growth parameters, the mean distance decreased in magnitude to -0.3712 mm (Fig. 6b). Similar convergence behaviors were observed for the other six patients.

**Fig 6.**
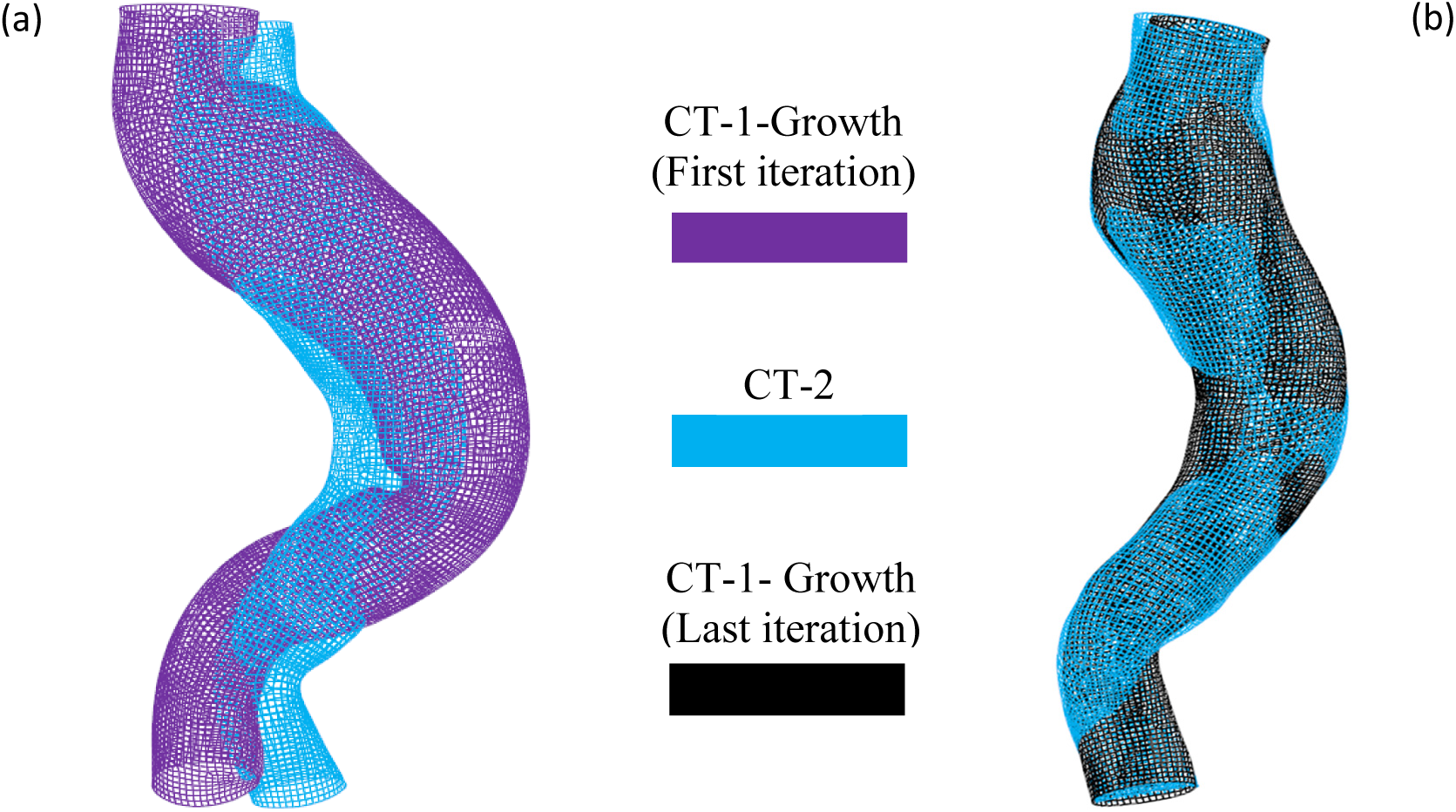
(a) Comparison of the FE-predicted CT-2 geometry (growth from CT-1, denoted CT-1-Growth) at the first iteration with the *in-vivo* CT-2 geometry for patient P1. (b) Comparison of the FE-predicted CT-2 geometry (CT-1-Growth) at the final iteration with CT-2.

For the seven patients (P1-P7), the mean node-to-surface distance between the *in-vivo* inner surfaces at CT-1 and CT-2 ranged from -3.7125 mm to -1.6042 mm (Table 4). With optimized growth parameters (Table 5), the mean distance between the FE-predicted CT-2 inner surface (growth from CT-1) and the *in-vivo* CT-2 geometry ranged from -0.8597 mm to +0.1648 mm.

**Table 4.**
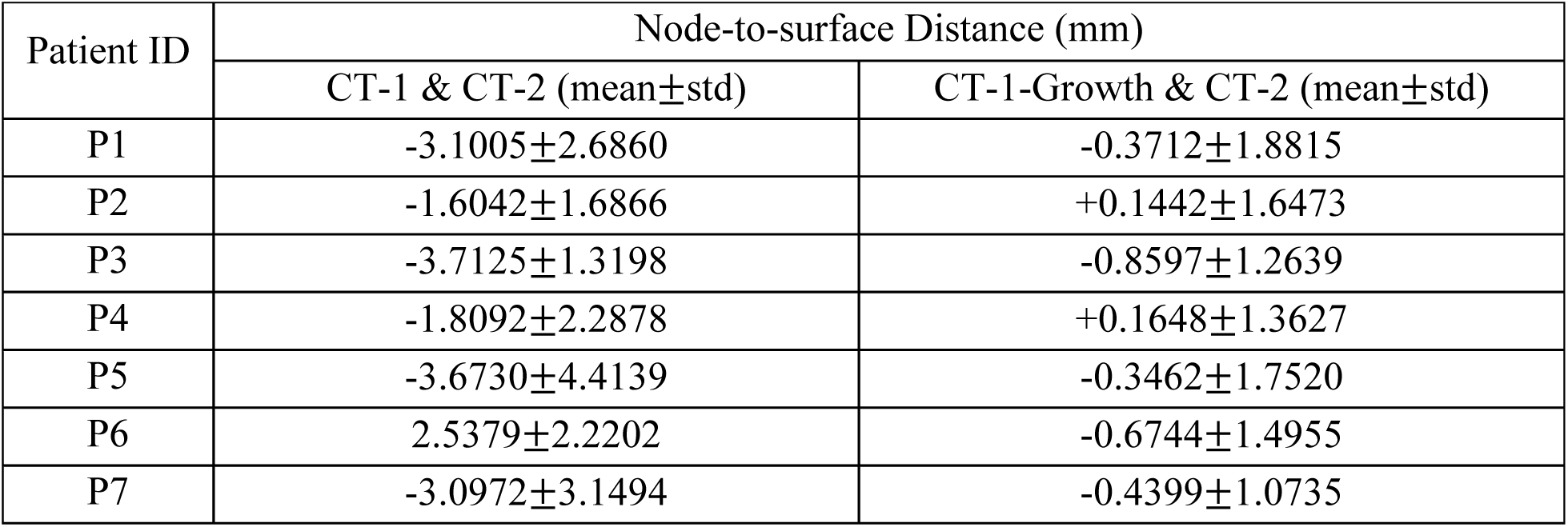
Mean and standard deviation (mean±std) of the node-to-surface distance between the *in-vivo* CT-1 and CT-2 inner surfaces for the descending aorta of seven patients (P1–P7), and between the FE-predicted CT-2 geometry grown from CT-1 (CT-1-Growth) and the *in-vivo* CT-2 geometry. The corresponding optimized growth parameters are listed in Table 5.

| Patient ID | Node-to-surface Distance (mm) |  |
| --- | --- | --- |
| | CT-1 & CT-2 (mean±std) | CT-1-Growth & CT-2 (mean±std) |
| P1 | -3.1005±2.6860 | -0.3712±1.8815 |
| P2 | -1.6042±1.6866 | +0.1442±1.6473 |
| P3 | -3.7125±1.3198 | -0.8597±1.2639 |
| P4 | -1.8092±2.2878 | +0.1648±1.3627 |
| P5 | -3.6730±4.4139 | -0.3462±1.7520 |
| P6 | 2.5379±2.2202 | -0.6744±1.4955 |
| P7 | -3.0972±3.1494 | -0.4399±1.0735 |

**Table 5.** Optimized growth parameters in Eq. (9) for the descending aorta of seven patients (P1–P7), corresponding to the CT-2 predictions obtained from growth of CT-1 (CT-1-Growth) reported in Table 4.

| | $\alpha_1$ (1/[MPa · Year]) | $\alpha_2$(1/[MPa · Year]) | $\sigma_{1c}$ (kPa) | $\sigma_{2c}$(kPa) |
| --- | --- | --- | --- | --- |
| P1 | 0.0576 | 0.0124 | 1.7557 | 4.5953 |
| P2 | 0.1624 | 0.1383 | 3.5090 | 4.3957 |
| P3 | 0.4697 | 0.1693 | 0.9184 | 0.5379 |
| P4 | 0.0362 | 0.0521 | 7.3914 | 0.6838 |
| P5 | 0.2519 | 0.2380 | 0.1235 | 0.1001 |
| P6 | 0.0259 | 0.0243 | 0.7425 | 0.1003 |
| P7 | 0.2207 | 0.2184 | 0.8172 | 0.5155 |

### 3.2 Growth Prediction Based on Optimized Growth Parameters

Across patients P1–P7, growth from CT-2 to CT-3 yielded mean node-to-surface distances ranging from -2.6500 mm to +3.0574 mm (Table 6). Using the optimized growth parameters in Table 5, the comparison between the predicted CT-3 geometry (CT-2-Growth) and the *in-vivo* CT-3 geometry showed good agreement, with mean node-to-surface distances ranging from -0.6651 mm to +0.3150 mm (Table 6). As an example, prediction results for patient P1 are shown in Fig. 7. To mitigate boundary effects, nodes belonging to the five-layer grid at the boundary of the upper and lower sections were excluded from analysis.

**Fig 7.**
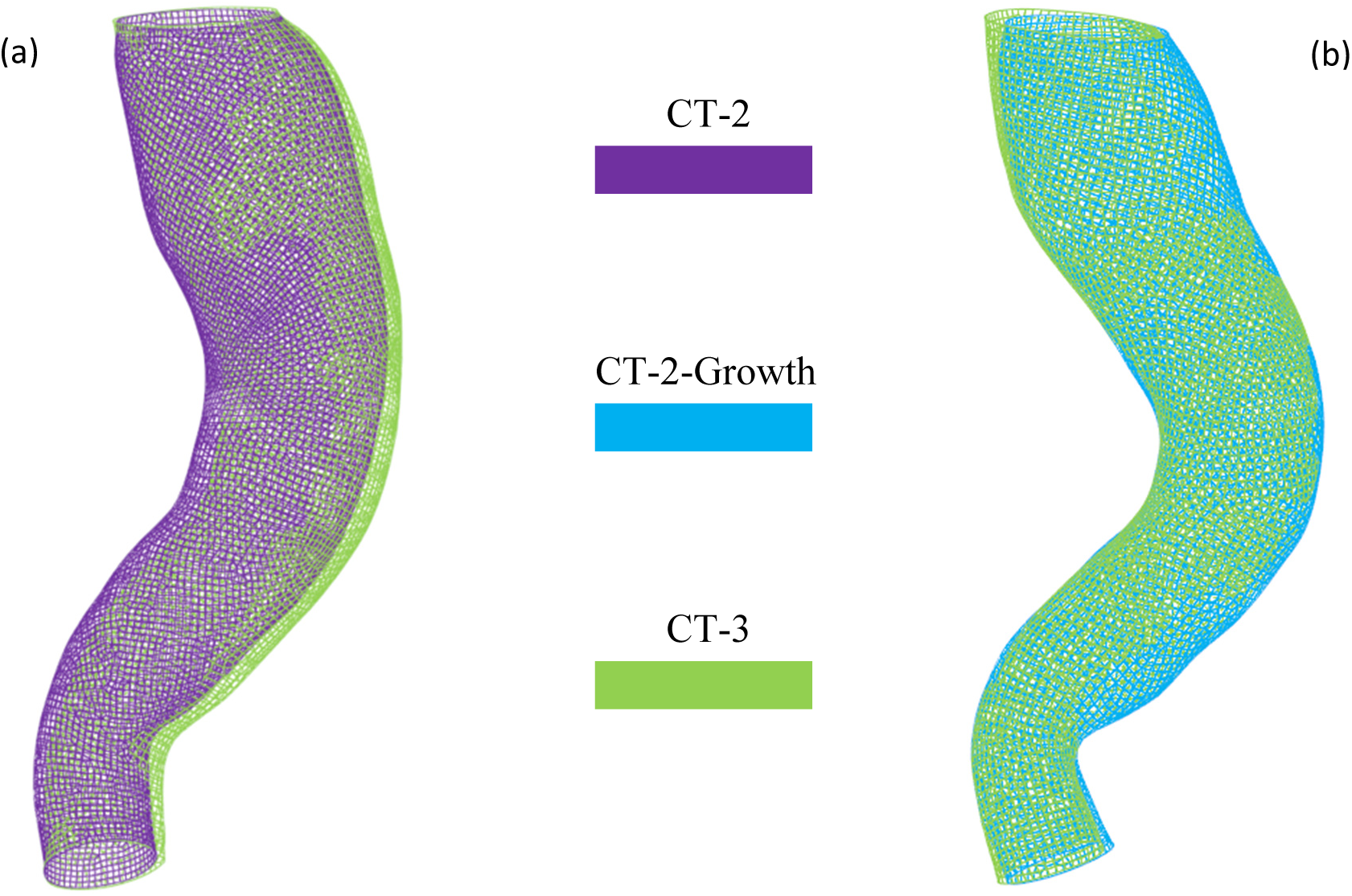
(a) Comparison of the *in-vivo* CT-2 and CT-3 geometries for the descending aorta of patient P1; (b) comparison of the predicted CT-3 geometry (growth from CT-2, CT-2-Growth) with the *in-vivo* CT-3 geometry.

**Table 6.** Mean and standard deviation (mean±std) of the node-to-surface distance between the *in-vivo* CT-2 and CT-3 inner surfaces for the descending aorta of seven patients (P1–P7), and between the predicted CT-3 geometry obtained by growth from CT-2 (CT-2-Growth) and the *in-vivo* CT-3 geometry.

| Patient ID | Node-to-surface Distance (mm) |  |
| --- | --- | --- |
|  | CT-2 & CT-3 (mean±std) | CT-2-Growth & CT-3 (mean±std) |
| P1 | -0.9612±1.9466 | +0.0479±0.9944 |
| P2 | -0.8284±1.1042 | -0.4062±0.9477 |
| P3 | -1.6146±1.1323 | -0.6651±0.9050 |
| P4 | -2.2214±1.4450 | -0.3441±0.7654 |
| P5 | +3.0574±1.6280 | +0.3150±1.8720 |
| P6 | -1.3027±1.9381 | -0.6113±1.5744 |
| P7 | -2.6500±1.6402 | -0.0028±0.4588 |

### 3.3 Diameter Prediction of Descending Aorta

We computed descending-aorta diameters for patients P1–P7 from the predicted CT-3 geometry (CT-2-Growth), which agreed well with the ground-truth CT-3 values (Fig. 8). For all seven patients, the maximum diameter error was <3.5%, and the mean diameter error was <4%. The mean ± standard error for the maximum and mean diameter errors were 2.05% ± 0.35% and 1.79% ± 0.48%, respectively. These results support the accuracy of our growth-based model for predicting descending-aorta diameter.

**Fig 8.**
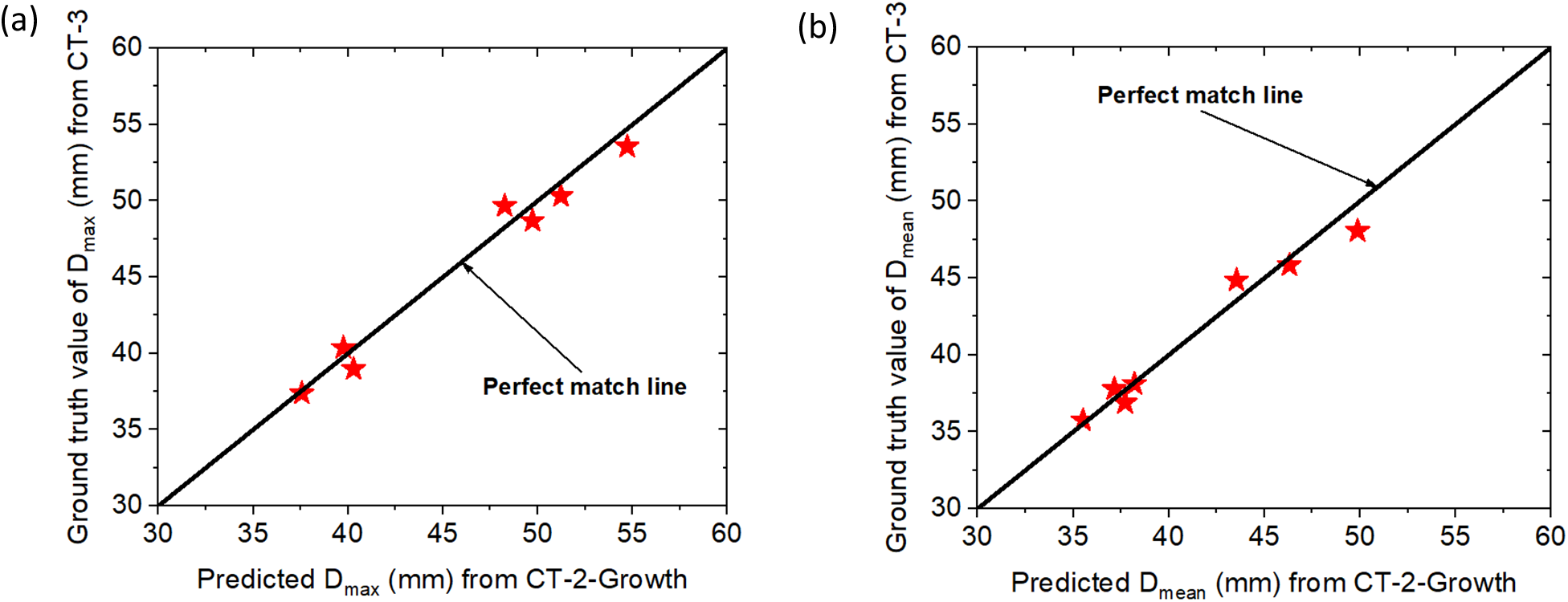
Diameter comparison between the predicted CT-3 geometry (CT-2-Growth) and CT-3 ground truth for seven patients (P1–P7): (a) D_max_(maximum diameter); (b) D_mean_(mean diameter).

## 4. Discussions

This study demonstrates that a wall stress-driven, UFD-based finite element (FE) model can accurately predict aortic growth in patients with uncomplicated type B aortic dissection (TBAD) under medical management. Using patient-specific geometry and blood pressure data, the model forecasted the false lumen aneurysmal expansion between follow-up scans with high fidelity: across seven patients, the predicted aortic geometry at the third time point closely matched the actual CT, with maximum diameter errors under 3.5% and mean errors under 4%. Such performance is notable given the variability in patient anatomies and follow-up intervals. The results suggest that this biomechanics-driven framework captures the essential aspects of TBAD enlargement, yielding patient-specific growth forecasts that could support clinical decision-making. Importantly, the model did not focus on individual idiosyncrasies of each case; instead, general patterns emerged. For instance, regions experiencing higher wall stress tended to exhibit greater outward growth, a trend consistent across the cohort. Even patients with relatively modest expansion were predicted well, indicating no obvious bias toward over-predicting or under-predicting growth. Overall, the UFD-based FE approach proved robust in reproducing both the magnitude and distribution of aortic enlargement in these seven cases.

Compared to other TBAD growth prediction approaches, this stress-driven modeling offers a distinctly different paradigm. Most prior computational studies of TBAD have emphasized hemodynamics rather than wall remodeling. For example, image-based computational fluid dynamics (CFD) simulations and 4D flow MRI analyses have been used to identify complex flow patterns that correlate with false lumen expansion[1, 37, 38]. These hemodynamic studies highlight important risk factors; indeed, flow metrics have been statistically linked to regions of aneurysmal growth. However, they generally stop short of predicting future geometry: a CFD or 4D-flow analysis might flag a patient as high-risk due to adverse flow conditions, but it will not quantitatively forecast how the aorta’s size will change over time. Similarly, fluid–structure interaction (FSI) models of TBAD have been developed to explore phenomena like dynamic flap motion, false lumen pressurization, and thrombus deposition[10, 39]. While FSI simulations provide insight into the mechanistic interplay between blood flow and wall mechanics, they too have not been used to project long-term vessel remodeling. In contrast, our FE growth model directly tackles the evolution of the vessel itself by incorporating a growth law – thus filling a gap left by pure fluid-based analyses.

A key strength of the stress-driven growth model is its potential for individualized prognosis. By accounting for each patient’s unique aortic geometry and blood pressure, the model inherently tailors the prediction to that patient. This is particularly valuable clinically because TBAD progression is notoriously heterogeneous. Some patients’ false lumens remain stable for years, while others expand rapidly despite similar initial presentations. Current clinical practice during optimal medical therapy (OMT) relies on serial imaging to catch those expansions. Studies have shown that even under medical management, a large fraction of TBAD patients will experience aneurysmal degeneration over time [4, 40], and even moderate growth correlates with heightened rupture risk. Consequently, surveillance protocols are aggressive, and elective intervention is often recommended if the aorta reaches certain triggers (e.g., a diameter beyond ∼5.5 cm or rapid growth exceeding ∼5.0 mm/year [41]). The ability to predict false lumen expansion during OMT could revolutionize this paradigm. Rather than reacting to anatomic changes after they occur, clinicians could anticipate them. For instance, our model could identify a patient whose dissected aorta, though currently below an intervention threshold, is on track to expand to dangerous dimensions within the next 6–12 months. Armed with that forecast, a surgeon might opt for earlier elective TEVAR – timed at a moment when the patient is still stable – instead of waiting until the aneurysm becomes urgent or emergent. In this way, a growth prediction tool can serve as a decision support system, aiding in the timing of therapy to prevent disaster. It is important to note, however, that any such model must be well validated; false positives (predicting growth that doesn’t occur) could lead to unnecessary procedures, while false negatives could give false reassurance. In our limited series, the model’s predictions were quite accurate, suggesting it has promise to reliably flag true expansion while avoiding false alarms.

Despite the encouraging performance, our modeling framework has several limitations that must be acknowledged. First, we assumed a uniform wall thickness for all patients, due to the lack of patient-specific thickness information from standard CT scans. In reality, dissection flaps and aortic walls can have variable thickness, and chronic dissection can involve scar tissue or calcifications that alter wall structure. This simplification could introduce error in stress calculations, since wall stress is inversely related to thickness. If, for example, a patient’s aortic wall is thinner than assumed in a certain region, the actual wall stress would be higher than our model predicts, potentially leading to underestimation of growth in that region. Second, the boundary conditions and loading in our simulation were simplified. We applied pressure to the aortic wall based on brachial blood pressure, and did not simulate the pulsatile nature of blood flow or any pressure differential between the true and false lumens. The model effectively treats the dissection as a single-wall structure experiencing average pressure, without capturing transient pressure spikes, flow-induced wall shear stress, or interactions with branching vessels and surrounding support. This absence of hemodynamic coupling means phenomena like flow oscillation, turbulence, or thrombus formation (which could alter pressure distributions) were neglected. Such factors are known to influence dissection progression, so excluding them limits the model’s completeness. Model validation on only seven patients is another limitation; this sample size, while providing initial evidence of feasibility, is too small to capture the full variability in TBAD presentations.

Looking forward, there are several directions for future work that could address these limitations and enhance the modeling framework. One important step is to expand the patient cohort to include more cases and a broader spectrum of TBAD anatomies. A larger study would help verify the model’s generalizability and might enable stratification of results by subgroups (for example, fully patent false lumen vs. partially thrombosed, or acute vs. subacute presentations). Another key avenue is integrating hemodynamics into the growth simulation. This could involve coupling the FE model with CFD or adopting a fluid–structure interaction approach, so that spatial maps of wall shear stress, true-false lumen pressure differences, and flow patterns inform the growth law. Incorporating such hemodynamic factors might capture aspects of TBAD progression that a wall-only model misses, potentially improving accuracy in cases where, say, an eccentric jet in the false lumen drives localized dilatation. Along similar lines, using time-varying pressure inputs (instead of a static pressure) in the FE simulation could account for the cyclic loading of the cardiac cycle. A pulsatile growth simulation would permit phenomena like stress fatigue or time-averaged growth effects to emerge naturally and might better represent the mechanobiological environment of the aorta over long periods. Beyond physics-based enhancements, there is an opportunity to leverage machine learning to augment the growth model. For example, ML algorithms could be trained on a database of simulated outcomes to predict patient-specific growth parameters from a single snapshot in time. A model could learn to infer the underlying tissue responsiveness (the calibrated parameters) from features of the aorta’s initial shape, false lumen size, or clinical indices. Additionally, ML could assist in real-time forecasting by providing faster surrogate models of the FE simulations, enabling clinicians to get near-instantaneous predictions after image acquisition. Finally, improvements in the biomechanical model itself are worth exploring. This includes allowing the wall thickness to evolve (or using patient-specific thickness estimates from advanced imaging), introducing nonlinear or multi-factor growth laws (e.g. incorporating both stress and stretch or biochemical cues), and accounting for tear evolution or collagen fiber remodeling as the dissection matures. By pursuing these future directions, the presented framework can be refined into a more comprehensive tool. In summary, while our current UFD-based FE model is an encouraging proof-of-concept for individualized TBAD growth prediction, ongoing enhancements and larger studies will be pivotal to translate it into a reliable clinical predictive instrument.

## 5. Conclusions

In this study, we presented a wall stress–driven, UFD model–based finite-element framework to predict post-dissection growth of the descending aorta in patients with uncomplicated type B aortic dissection. Using three serial CT scans from seven patients, a centerline-based algorithm and inverse optimization of growth parameters enabled patient-specific simulations that reproduced both the magnitude and spatial distribution of aortic enlargement over two follow-up intervals with small geometric and diameter errors. The strong agreement between predicted and observed diameters at the third CT suggests that stress-regulated growth laws can capture the underlying mechanobiology of aortic remodeling after dissection. This computational framework therefore provides a promising foundation for digital-twin models that may ultimately improve individualized surveillance, refine thresholds for thoracic endovascular aortic repair, and reduce unnecessary interventions. Future work may include prospective validation in larger cohorts and incorporate additional factors such as intramural thrombus, wall heterogeneity, evolving hemodynamics, and dynamic loading effects over time.

## Data Availability

All data produced in the present study are available upon reasonable request to the authors

## Acknowledgment

This study is supported by NIH R01HL155537 and grants from the Carlyle Fraser Heart Center. The author H.D. thanks Kristina Porte for her assistance with data collections.

## Appendix A: Resolution of CT Images

The in-plane spatial resolution of the CT images for the seven patients (P1–P7) ranged from 0.69 × 0.69 mm to 1.07 × 1.07 mm, with slice thicknesses of 1.0–2.0 mm. Detailed CT image resolutions for each patient are listed in Table A1.

**Table A1:** In-plane resolution and slice thickness of CT images at CT-1, CT-2, and CT-3 for the descending aorta in patients P1–P7.

| Patient ID | CT scan | In-plane resolution (mm) | Slice thickness (mm) |
| --- | --- | --- | --- |
| P1 | 1 | $0.87 \times 0.87$ | 1.25 |
| | 2 | $0.87 \times 0.87$ | 1.50 |
| | 3 | $0.98 \times 0.98$ | 1.50 |
| P2 | 1 | $0.71 \times 0.71$ | 1.00 |
| | 2 | $0.77 \times 0.77$ | 1.50 |
| | 3 | $0.80 \times 0.80$ | 1.50 |
| P3 | 1 | $1.07 \times 1.07$ | 1.07 |
| | 2 | $0.70 \times 0.70$ | 1.25 |
| | 3 | $0.70 \times 0.70$ | 1.25 |
| P4 | 1 | $0.80 \times 0.80$ | 1.25 |
| | 2 | $0.78 \times 0.78$ | 1.00 |
| | 3 | $0.98 \times 0.98$ | 1.50 |
| P5 | 1 | $0.77 \times 0.77$ | 2.00 |
| | 2 | $0.90 \times 0.90$ | 1.25 |
| | 3 | $0.91 \times 0.91$ | 1.50 |
| P6 | 1 | $0.69 \times 0.69$ | 1.50 |
| | 2 | $0.82 \times 0.82$ | 1.00 |
| | 3 | $0.86 \times 0.86$ | 1.50 |
| P7 | 1 | $0.72 \times 0.72$ | 1.00 |
| | 2 | $0.71 \times 0.71$ | 1.25 |
| | 3 | $0.75 \times 0.75$ | 1.50 |

